# popexposure: An open-source Python package to find the number of people residing near environmental hazards quickly and efficiently

**DOI:** 10.1101/2025.10.19.25338326

**Authors:** Heather McBrien, Joan A. Casey, Lawrence G. Chillrud, Nina M. Flores, Lauren B. Wilner

## Abstract

Environmental scientists often assess exposure to hazards using residential proximity (i.e., they consider an individual living near a hazard to be exposed). Such assessment requires large, fine-scale spatial datasets that describe locations of environmental hazards and residential populations. Manipulating such datasets is technically demanding, slow, memory-intensive, and difficult to optimize for speed and memory use. Currently, individual research teams each write their own algorithms for this task. This may lead to inconsistencies in assumptions, methods, and results.

We developed an open-source Python package, *popexposure*, which quickly, efficiently, and consistently estimates the number of people living near environmental hazards. Given a set of distinct hazard geometries and corresponding buffer distances, *popexposure* can estimate the number of people living within the buffered area of each hazard using a gridded population dataset. *popexposure* can also estimate the number of people living within the buffer distance of each hazard by additional administrative geographies. For example, users can calculate the number of people exposed to hazards in each census tract or zip code tabulation area (ZCTA). *popexposure* addresses common issues encountered in this calculation: whether or not to double-count people exposed to more than one hazard, proper pixel apportionment, choosing appropriate map projections for data covering large areas, and optimizing speed and memory.

In this paper, we describe *popexposure*’s functionality and provide an example use case, calculating the proportion of people exposed to any wildfire burn zone disaster in California in 2018 in each ZCTA.

**What this study adds:** Environmental epidemiologists often assess exposure to hazards using residential proximity (i.e., they consider an individual exposed if they live near a hazard). This computation presents technical difficulties, and different research teams each apply their own solution, since no software currently exists to do this task. We developed an open-source Python package, *popexposure*, which quickly, efficiently, and consistently estimates the number of people living near environmental hazards. Here, we describe the package and provide an example use case, applying *popexposure* to compute the proportion of people exposed to any wildfire burn zone disaster in California in 2018 in each ZCTA.

## Intro

To study the relationship between environmental exposures and health outcomes, environmental epidemiologists often assess exposure using proximity (i.e., they consider an individual living near a hazard as exposed). Using this method, studies have assessed population exposure to wildfire disasters (1), oil and gas wells (2,3), Superfund sites (4), industrial emissions sources (5,6), tropical cyclones (7), and more. This method requires hazard data: geospatial data that contain polygons of hazard boundaries (e.g., wildfire burn zones), lines of hazard paths (e.g., tropical cyclone tracks), or points of hazard locations (e.g., gas well coordinates). To find populations exposed to these hazards, researchers may buffer hazards by an exposure- or health-relevant distance and then overlay the geospatial hazard data with a gridded population raster to determine the number of people living near the environmental hazards.

This computation is technically demanding. When an analysis involves large, fine-scale spatial datasets (e.g., exposure to oil and gas wells, which total millions of exposure points in the US) (8) or datasets that cover a large area (e.g., national or global analyses of exposure), overlaying hazard data with population data requires substantial computational power, to which all research teams may not have access. Currently, individual research teams independently develop and use algorithms for this task. This may lead to inconsistencies in assumptions, methods, and results. For example, exposed population estimates can vary widely depending on the raster masking procedure and how partial pixel intersections between the hazard and raster are handled.

We created an open-source Python package, *popexposure*, which quickly, efficiently, and consistently estimates the number of people living near environmental hazards. It is optimized for speed and memory use and can be installed with *pip*. In this paper, we describe the class and methods available in *popexposure* and provide an example of its use. We find the proportion of people exposed to any wildfire burn zone disaster in California in 2018 in each zip code tabulation area (ZCTA).

## Methods

### Package functionality

*popexposure* identifies the number of people living near environmental hazards by first buffering the geospatial environmental hazard data and then overlaying it with a gridded residential population raster. Then, it sums the raster values in the buffered hazard areas to find the number of residents exposed.

In summary, to estimate the number of people living near environmental hazards, *popexposure* takes up to four inputs: 1: a geospatial dataset of environmental hazards, 2: a gridded population dataset, 3: buffer distances around those hazards, and 4: an optional geospatial dataset of administrative units such as census tracts, ZCTAs, or counties. *popexposure* can calculate either a single count of people living within one or more buffer zones of any hazards in the dataset, or separate counts for each unique hazard, estimating how many people live within the buffer zones of each individual hazard. These exposure estimates can also be broken down by administrative units such as ZCTAs; for example, *popexposure* can find the number of people living within 10 km of one or more US wildfire disaster burn zones in 2018 by ZCTA.

Some common problems arise when trying to compute counts of residential populations exposed to environmental hazards. They include 1: overlapping hazard or buffered hazard geometries and how to count people who reside within two or more geometries; 2: selecting a buffer distance based on an attribute of the hazard (e.g., more reported sulfur dioxide emissions from coal plants could merit a larger buffer distance); 3: using appropriate map projections when analysis spans large geographies; 4: apportionment of people identified in partially exposed pixels; and 5: challenges with memory use and computation speed. *popexposure* offers standardized solutions to each problem, described below in the key features section.

### Key features

1. <u>Counting people exposed to more than one hazard</u>: In a set of environmental hazards, the hazards or their affected areas may overlap. *popexposure* allows users to choose whether to estimate exposure to each hazard individually or to the entire group of hazards combined. Users can calculate a single count representing the number of people exposed to one or more hazards in the combined area of all hazards, or separate counts for each unique hazard, showing how many people live near each specific hazard.
2. <u>Buffer based on a hazard attribute</u>: Users can specify different buffer distances for each hazard by varying the buffer distance value by hazard. Users can also provide multiple buffer distances per hazard by including one or more buffer distance columns in the data. Users may also do both. These distances can be the same for all hazards or vary depending on characteristics of each hazard, such as type, size, or severity. For example, larger wildfires could have larger buffers based on their burned area. To analyze exposure at multiple distances—such as 10 km and 20 km—users can include multiple columns, each representing a different distance for every hazard.
3. <u>Map projections</u>: The user may pass data with any coordinate reference system to *popexposure*, and the package will use the buffer distances created and passed by the user to buffer the hazard geometries in the best Universal Transverse Mercator (UTM) projections for each set of environmental hazards (9), based on each hazard’s centroid latitude and longitude, minimizing map distortion. For example, for a hazard dataset spanning the continental United States, each individual hazard would be reprojected for buffering into one of the 10 UTM projections spanning the United States based on its centroid latitude and longitude.
4. <u>Population apportionment</u>: *popexposure* masks the gridded population raster with the buffered hazard geometries using partial pixel masking, using the package *exactextract* (10). This means if the hazard geometry overlaps with half of pixel *i* which contains 100 people then when all pixel values within the buffered hazard geometry are summed, only 50 people from pixel *i* will be added to the sum of exposed individuals. This approach assumes an even population distribution across the pixel but produces the most accurate count of people affected, in contrast to centroid masking or including the entire value of any pixels touched by the hazard geometry.
5. <u>Computational challenges</u>: Raster masking is done using *exactextract* for each individual geometry or set of overlapping geometries in the set of hazards to minimize working memory use and maximize computation speed. *exactextract* implements a fast algorithm^1^ by avoiding point-in-polygon tests to determine which raster cells intersect with polygons, outperforming similar packages.

### Available functions

The package contains one main class, with two methods.

#### 1. Main class: PopEstimator

This class contains two methods. `est_exposed_pop()` finds the number of people living near hazards, and `est_total_pop()` finds the number of people living in administrative units. The `PopEstimator` class has two attributes, which are both optional for initialization: population data (`pop_data`), and optional administrative unit data (`admin_data`). Population data do not require any preprocessing, if a path to these population data is passed, it is stored as the `pop_data` attribute.

When `admin_data` are passed, the `PopEstimator` reads, cleans, and preprocesses the data to prepare them for input into the `est_exposed_pop()` and `est_total_pop()` methods. It loads a geospatial file (GeoJSON or GeoParquet) containing administrative geographies (e.g., ZCTAs, census tracts). It ensures that all geometries in both files are valid and removes any that are empty, missing, or impossible to make valid. Rows with empty or missing geometries are removed, and their corresponding hazard or administrative unit IDs are removed from the dataset stored in the `PopEstimator`. If the input file is empty or contains no valid geometries, the `admin_data` attribute is set to `None`. This is also true if no administrative unit data are passed.

### Initialization inputs

i. `pop_path`: A string path to a gridded population raster file, in TIFF format. The raster may have any coordinate reference system (optional).
ii. `admin_data`: A string path to a geospatial data file (.`geojson` or `.parquet`) containing administrative geography data. Data may have any coordinate reference system. Administrative unit data must contain a string column `ID_admin_unit` with unique admin unit IDs and a geometry column `geometry` (optional).

#### 2. est_exposed_pop()

`est_exposed_pop()` estimates the number of people living within specified distances of environmental hazards using a gridded population raster. If population data were loaded when a `PopEstimator` was initialized, it uses these data. Otherwise, the user must pass population data. If a `PopEstimator` was initialized with administrative units, it uses those, as well as hazard data. This method can calculate exposure either separately for each hazard (hazard-specific), producing one count of people exposed per hazard, or for the combined area of all hazards together (cumulative), producing a single overall count of anyone exposed to one or more hazards in the set. When administrative units are provided, the population counts are calculated within the intersection of each buffered hazard geometry and each administrative unit. For partial pixel overlaps, population counts are weighted proportionally by the area of intersection. The method can handle multiple buffer distances simultaneously, producing exposure estimates for each specified buffer.

Hazard data must include one or more user-specified numeric columns that define buffer distances in meters. These buffer distances determine how far a person must live from each hazard to be considered exposed. For example, to identify people living within 10 km of each hazard, the user should include a column with all values set to 10,000 (representing 10,000 m because the UTM projection uses meters). To calculate exposure for multiple buffer zones (e.g., 10 km and 20 km), the user should include two columns: one with all values set to 10,000 and another with all values set to 20,000. Each buffer column must have a name starting with “`buffer_dist`” and end with a unique suffix (e.g., e.g., “`buffer_dist_main`”, “`buffer_dist_sensitivity`”). To apply different buffer distances to different hazards, the user should enter varying values within any single buffer distance column, so that each hazard has its own specific buffer distance within that column. For example, the user could apply a larger buffer distance to wildfires that burned greater than a certain acreage.

For each hazard, `est_exposed_pop()` creates buffered geometries using the best-fitting UTM projection for accurate distance-based operations. It then reprojects the buffered geometries back to a common coordinate reference system.

Users can choose to estimate either (a) hazard-specific counts: the number of people exposed to each unique buffered hazard. For example, for hazard-specific counts and an input set of 10 distinct hazards, `est_exposed_pop()` will output 10 distinct counts of people exposed: one for each hazard. In this case, people exposed to each hazard are not mutually exclusive and may be double-counted or more. Users may also estimate (b) a cumulative count: the number of unique people exposed to the combined affected area of all input hazards. For example, for the same input set of 10 distinct hazards, `est_exposed_pop()` can output a single estimate of the number of unique people exposed to one or more of the 10 input buffered hazards, thereby avoiding “double counting” people exposed to more than one hazard in the input set.

Either type of estimate can be broken down by additional administrative geographies such as ZCTAs; for example, the user can estimate the number of people within 10 km of each unique wildfire burn zone disaster in 2018 by ZCTA. Users must supply at least one buffered hazard column in the input data, but may supply additional buffered hazard columns to create estimates of exposure for different buffer distances.

### Inputs

i. `hazards`: A `GeoDataFrame` with a coordinate reference system containing a string column `ID_hazard` with unique hazard IDs, and one or more geometry columns starting with `buffered_hazard` containing buffered hazard geometries. “`buffered_hazard`” columns must each have a unique suffix (e.g. columns could be “`buffered_hazard_main`”, “`buffered_hazard_sensitivity`”, and “`buffered_hazard_large_buffer`”).
ii. `hazard_specific`: A `Boolean (True/False)` indicating if exposure should be calculated for each hazard individually (hazard-specific estimates), or if geometries should be combined before exposure is calculated, producing a single cumulative estimate.
iii. `pop_path`: A string path to a gridded population raster file, in TIFF format. The raster may have any coordinate reference system. (Population data are not required here if they were already provided when initializing the estimator).

### Outputs

A `DataFrame` with the following columns:

- `ID_hazard`: Always included.
- `ID_admin_unit`: Included only if admin units were provided.
- One or more “`exposed`” columns: Each corresponds to a buffered hazard column (e.g., if the input had columns “`buffered_hazard_main`”, “`buffered_hazard_sensitivity`”, and “`buffered_hazard_large_buffer`”, the output will have “`exposed_main`”, “exposed_sensitivity”, and “`exposed_large_buffer`”). Each “`exposed`” column contains the sum of raster values (population) within the relevant buffered hazard geometry or buffered hazard geometry and administrative unit intersection.

The number of rows in the output `DataFrame` depends on the method arguments:

- If `hazard_specific` was True, the `DataFrame` contains one row per hazard or per hazard-administrative unit pair, if administrative units are provided.
- If `hazard_specific` was `False`, the `DataFrame` contains a single row or one row per administrative units, if administrative units are provided, with each “`exposed`” column representing the total population in the union of all buffered hazard geometries in that buffered hazard column. E.g., the column “`exposed_10`” in the method output would indicate the number of people exposed to any buffered hazard in “`buffered_hazard_10`”.

There are four ways to use this method:

1. **Hazard-specific exposure, no additional administrative geographies** (`hazard_specific` = `True, admin_units` = `None`): Calculates the exposed population for each buffered hazard geometry. The method returns a `DataFrame` with one row per hazard and one “`exposed`” column per buffered hazard column. If people lived within the buffer distance of more than one hazard, they are included in the exposure counts for each hazard they are near.
2. **Combined hazards, no additional administrative geographies** (`hazard_specific` = `False, admin_units` = `None`): All buffered hazard geometries in each buffered hazard column are merged into a single geometry, and the method calculates the total exposed population for the union of those buffered hazards. The method returns a `DataFrame` with a single row and one ‘exposed’ column for each buffered hazard column. This means that if people were close to more than one hazard in the hazard set, they are counted once (i.e., not ‘double counted’).
3. **Hazard-specific exposure within administrative units** (`hazard_specific` = `True, admin_units are provided`): The method calculates the exposed population for each intersection of each buffered hazard geometry and each administrative unit. Returns a `DataFrame` with one row per buffered hazard - administrative unit pair and one “`exposed`” column per buffered hazard column. If people lived within the buffer distance of more than one hazard, they are included in the exposure counts for their administrative unit-hazard combination for each hazard they are near.
4. **Combined hazards within administrative units** (`hazard_specific` = `False, admin_units provided`): All buffered hazard geometries in the same column are merged into a single geometry. The method calculates the exposed population for the intersection of each buffered hazard combined geometry with each administrative unit. Returns a `DataFrame` with one row per administrative unit and one “`exposed`” column per buffered hazard column. If people were close to more than one hazard in the hazard set, they are counted once.

#### 3. est_total_pop()

This method estimates the total population living within administrative geographies (e.g., ZCTAs, census tracts) using a gridded population raster. (Administrative data are supplied to a supplied to a `PopEstimator` during initialization, and population data can be provided during initialization or directly to this method as an argument). It is designed to be used with the same `PopEstimator` object, and the same population raster as the `est_exposed_pop()` method, to provide consistent denominators for calculating exposure percentages within each administrative unit. Although official population estimates often exist for these geographies, this method uses the gridded population data to recalculate these population estimates ensure consistency with the exposure estimates. The method sums the raster population values that fall within the boundaries of each administrative geography, applying the same spatial intersection logic used in `est_exposed_pop()` to handle partial overlaps between raster pixels and polygons.

i. `pop_path`: Optional. A string path to a gridded population raster file, in TIFF format. The raster may have any coordinate reference system. (Population data are not required here if they were already provided when initializing the estimator).

### Outputs

i. A `DataFrame` with an `ID_admin_unit` column matching the input and a `population` column, where each value is the sum of raster values within the corresponding administrative unit geometry.

### Results: Example

We used *popexposure* to identify populations exposed to wildfire burn zone disasters using a national, publicly available dataset maintained by our team, described in detail in Gordon et al. 2025.(11) Briefly, we conducted a literature review and developed a definition of wildfire burn zone disasters, and then synthesized data from several national US wildfire burn zone datasets. Wildfire burn zones met our disaster criteria if they burned near a community (≥ 96 people per km^2^) and resulted in ≥ 1 civilian fatality, ≥ 1 destroyed structure, or received federal disaster relief.(12) In addition to other information, this dataset contains geometries of each wildfire burn zone disaster’s spatial extent and ignition and containment dates.

We created a demonstration, available on the *popexposure* GitHub page, which illustrates four different ways *popexposure* could be used to assess exposure to wildfire burn zone disasters. A Jupyter notebook containing a full demonstration of how to use *popexposure* is available here: https://github.com/heathermcb/popexposure in the tutorial directory, as well as on our website: https://heathermcb.github.io/popexposure/.

The example used *popexposure* to find the proportion of people exposed to any wildfire burn zone disaster in California in 2018 in each ZCTA. We also calculated the total number of people living within 10 km of any wildfire burn zone disaster in California in 2018.

To do this, we passed the following inputs to a `PopEstimator` object and then to `est_exposed_pop()`:

1. The 2020 Global Human Settlement 100-meter resolution gridded population dataset as a raster .tif file.(13)
2. A TIGER/Line shapefile containing 2020 US ZCTA boundaries downloaded from census.gov and saved as a GeoJSON file.(14)
3. The wildfire burn zone disaster dataset containing spatial extents in the United States 2000-2019, subsetted for this example to California in 2018 (**Figure 1**), as a GeoParquet file, with a column specifying a buffer distance of 10 km for each fire.(11)

**Figure 1.**
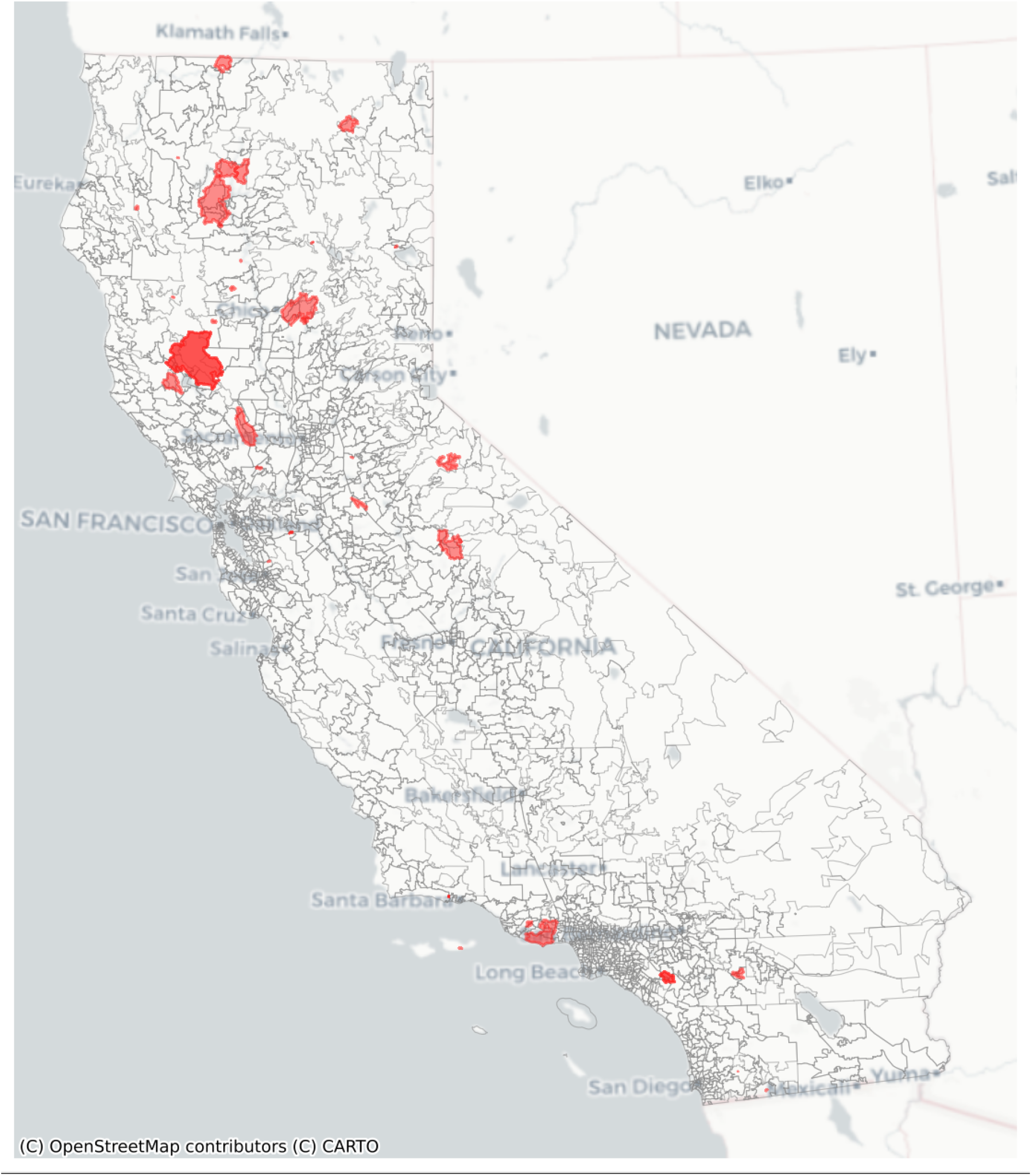
Wildfire burn zone disasters in California in 2018 overlaid on 2020 US census zip code tabulation areas.

Though ZCTA-level population data are available, we also used `est_total_pop()` to estimate the residential population of each ZCTA according to the GHSL dataset (**Figure 2**) for consistency between the numerator and denominator data in this example. **Figure 3** shows the head of example input and output datasets to `est_total_pop()`.

**Figure 2.**
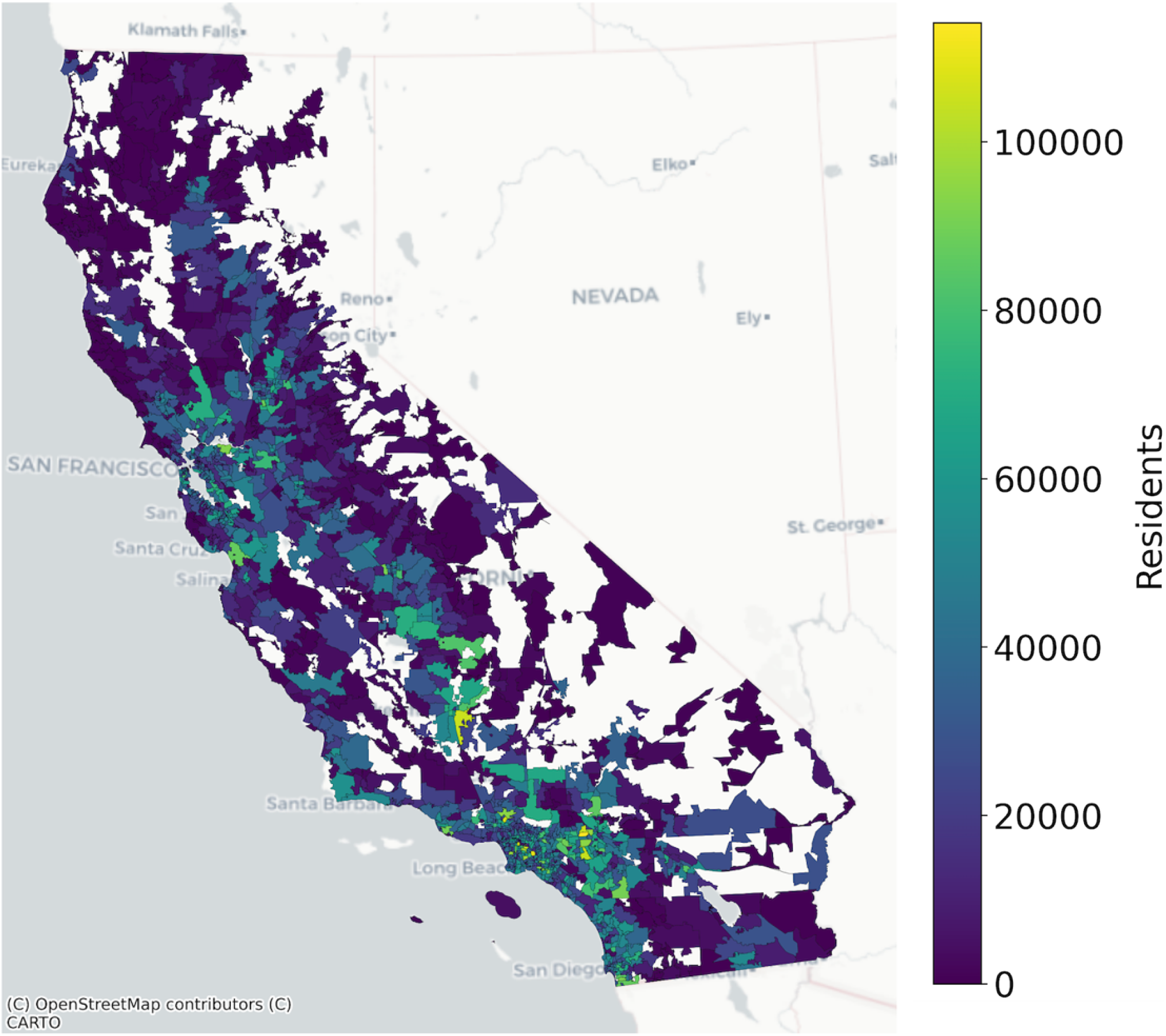
California 2020 zip code tabulation area population as estimated by the 2020 Global Human Settlement Layer 100m resolution gridded population dataset.

**Figure 3.**
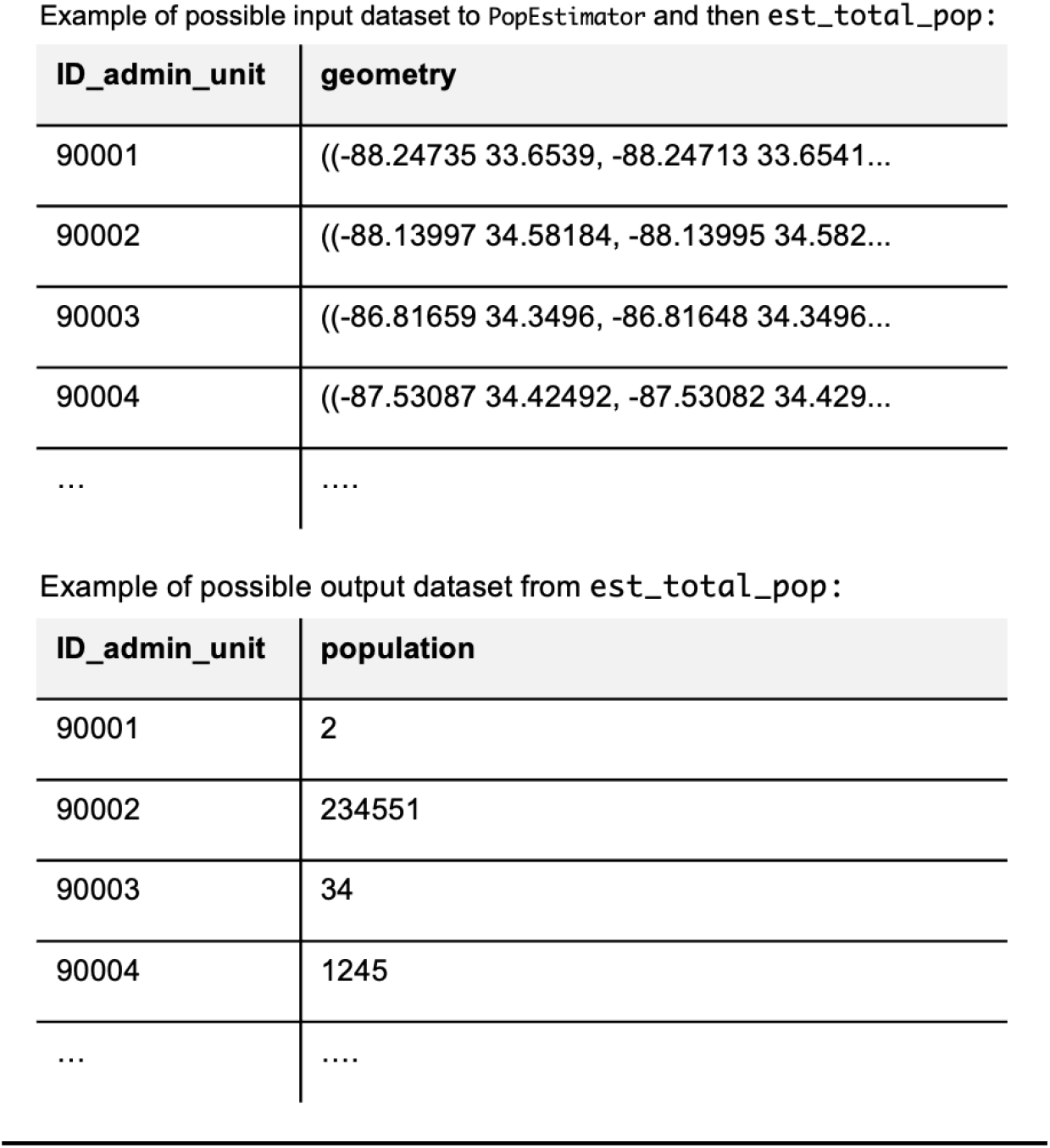
Head of example datasets that could be the input and output from `est_total_pop` in *popexposure*, which finds the number of people living in each geometry of the input dataset according to a gridded population raster file.

Then, we used `est_exposed_pop()` with `hazard_specific` = True to calculate the total number of people in each ZCTA living within the buffered boundaries of one or more fires (**Figure 4** shows examples of possible input and output datasets). We combined these outputs to find the proportion of people exposed to any wildfire burn zone disaster in California in 2018 in each ZCTA (**Figure 5**). The Woolsey Fire(15) (near Malibu) and the Camp Fire(16) (near Paradise) both burned during this year and affected large populations. Using *popexposure*, we found that 741,021 people lived within 10km of the Woolsey Fire or within its burn zone, and 157,271 people lived within 10km of the Camp Fire or within its burn zone. 5,320,181 people lived within 10 km of one or more wildfire burn zone disaster boundaries in 2018 in California.

**Figure 4.**
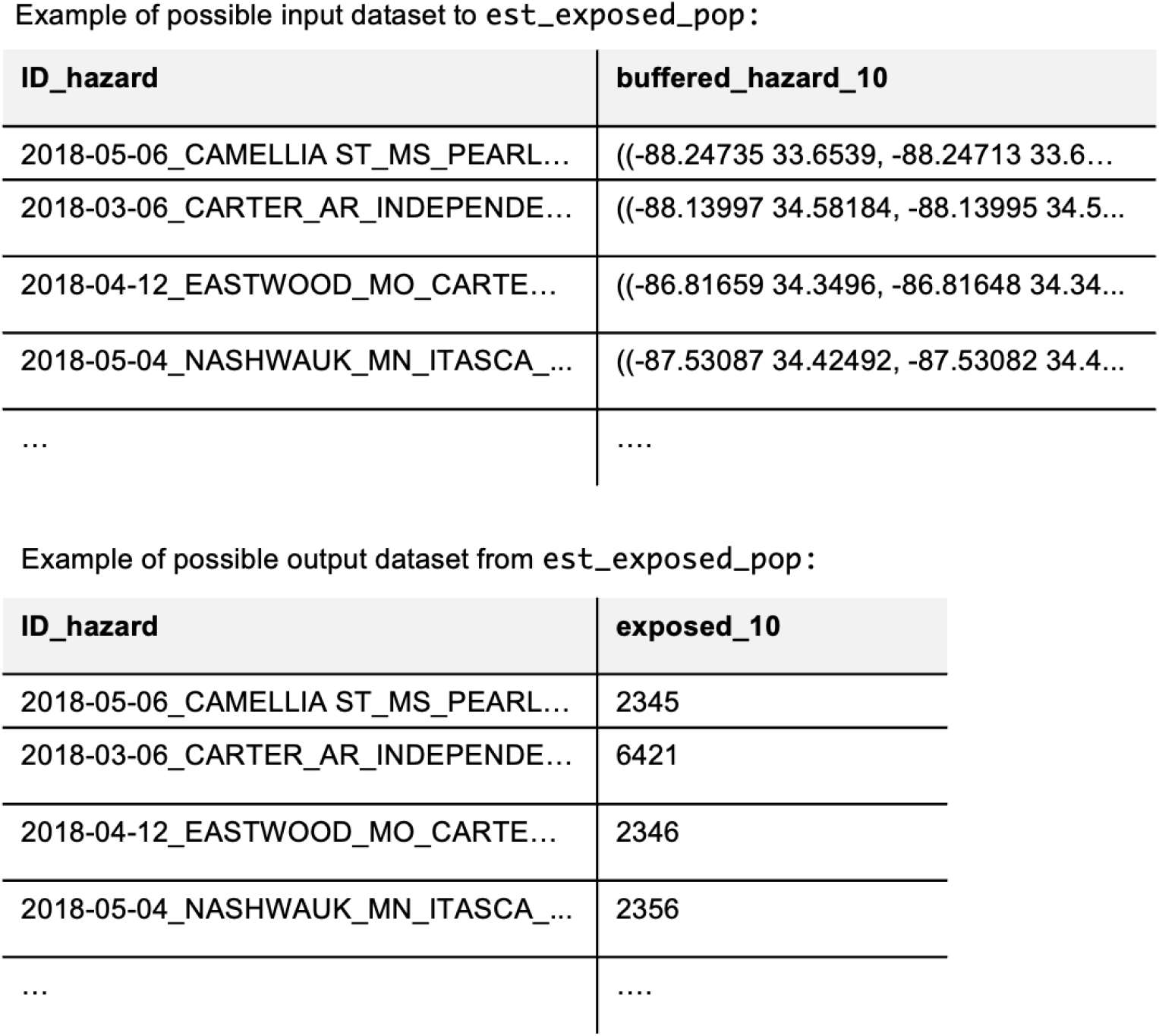
Head of example datasets that could be the input and output from `est_exposed_pop` in *popexposure*, which finds the number of people living within a buffer distance in each hazard geometry of the input dataset according to a gridded population raster file.

**Figure 5.**
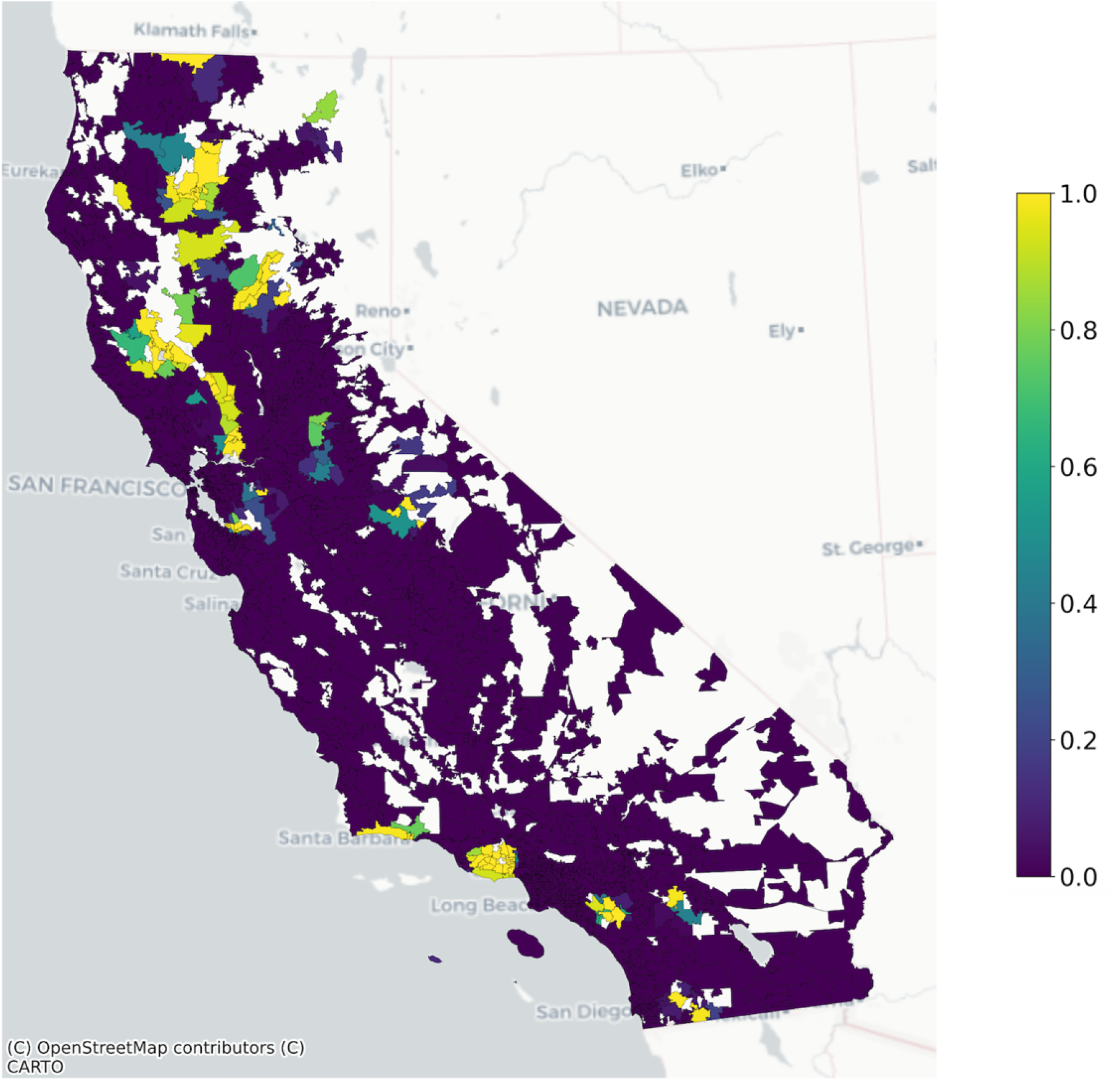
Proportion of the 2020 zip code tabulation area population living within 10 km of a wildfire burn zone disaster in California in 2018.

## Conclusion

Researchers have used different methods to accomplish a common task in environmental science: estimating the number of people living near environmental hazards. In the package *popexposure*, we offer a standardized and reproducible method for efficiently and accurately estimating people exposed to environmental hazards. In this paper’s example, *popexposure* found the number of people living near wildfire burn zone disasters in California in 2016, 2017, and 2018 in less than five seconds, and can find the number of people living near wildfire burn zone disasters in the United States from 2000-2020 in ten minutes on a laptop our research team had available for data analysis (2021 MacBook Pro with M1 Max chip and 64 GB RAM). We have included further benchmarking in our project GitHub repository. We hope the use of this package can streamline the common task of estimating populations exposed to environmental hazards, which has the potential to promote consistency and accuracy in exposure assessments broadly.

**<u>Note to reviewers: We have currently published a development version of the package, with the intent to release a stable version after peer review.</u>**

## Data Availability

All code and data are available at https://github.com/heathermcb/popexposure

## Ethics approval

This work does not involve human subjects, human material, or human data, therefore ethics approval was not required.

The authors have no conflicts of interest.

All code and data are available at: https://github.com/heathermcb/popexposure

## Funding sources

CIHR Doctoral Foreign Study Award (HM)

National Institute of Environmental Health Sciences (F31ES035280, NMF) National Institute on Aging (R01AG071024, JAC)

NSF GRFP DGE-2234667 (LGC)

## Footnotes

1 https://isciences.github.io/exactextract/background.html#algorithm

